# Blood epigenetic remodeling in asthma associated with biologic therapy

**DOI:** 10.64898/2026.09.24.26363868

**Authors:** Mario Martin-Almeida, Javier Perez-Garcia, Elena Martin-Gonzalez, Paloma Poza-Guedes, José Antonio Cañas, Inmaculada Sanchez-Machin, José María Hernández-Pérez, Daniel Gonzalez-Garcia, Elena Mederos-Luis, Berta Román-Bernal, Jesús Villar, Marcela Valverde-Monge, Diana Betancor, M^a^ Jesus Rodriguez-Nieto, Fabian Lorenzo-Diaz, Victoria del Pozo, Ruperto González-Pérez, Maria Pino-Yanes

**Author notes:** Corresponding author: Maria Pino-Yanes, Ph.D. Genomics and Health Group, Department of Biochemistry, Microbiology, Cell Biology, and Genetics, Universidad de La Laguna (ULL). Apartado 456, La Laguna, 38200 Santa Cruz de Tenerife, Tenerife, Spain.

## Abstract

**Background:** Polygenic risk scores (PRS) capture inherited susceptibility to asthma, whereas methylation risk scores (MRS) may reflect disease-associated epigenetic variation. We assessed whether these scores differ in patients requiring biologic therapy and whether MRS change after mepolizumab initiation.

**Methods:** Cross-sectional analyses in the GEMAS study included 699 participants with genotype data (428 with asthma, 271 controls) and 668 with blood DNA methylation data (408 with asthma, 260 controls). We evaluated 35 existing asthma PRS and derived six MRS from published epigenomic asthma studies. Longitudinal MRS changes after mepolizumab initiation were assessed in 16 GEMAS patients and were replicated in 20 patients from the MEGA study.

**Results:** Twenty-eight PRS differentiated biologic-treated patients from controls (areas under the curve [AUC], 0.68-0.75), but none distinguished biologic-treated and non-treated patients after multiple-testing correction. MRS was higher in asthma without biologic than in controls (odds ratio [OR], 1.64; 95% CI, 1.34-1.99) and lower in biologic-treated than in asthma without biologic patients (OR, 0.54; 95% CI, 0.35-0.81). MRS did not differ significantly between biologic-treated patients and controls. Following mepolizumab initiation, MRS decreased at 6 and 12 months in GEMAS (*p*=4.81×10^−5^ and *p*=2.40×10^−5^, respectively). The 6-month decrease was also observed in MEGA (*p*=3.25×10^−9^). MRS epigenetic markers were predominantly hypermethylated, opposite to their described pattern in asthma risk.

**Conclusions:** Asthma PRS did not differ significantly between biologic-treated and non-biologic-treated patients, but asthma MRS were lower in biologic-treated patients. The MRS decreased after mepolizumab initiation in two independent cohorts, consistent with a reversal of asthma-associated epigenetic patterns.

---

Polygenic risk scores (PRS) quantify inherited susceptibility to a trait, whereas methylation risk scores (MRS) integrate DNA methylation (DNAm) markers that may capture gene-environment interactions. PRS have shown moderate predictive performance for asthma, but whether genetic burden differs in patients requiring biologic therapy remains unclear^1^. Similarly, MRS derived from blood epigenome-wide association studies (EWAS) have not been evaluated in the context of biologic therapy. We aimed to assess the association of PRS and MRS of asthma with biologic-treated asthma.

We studied participants from the Genomics and Metagenomics of Asthma Severity (GEMAS) study. After quality control (**Tables S1-2**), genome-wide genotyping data were available for 699 participants (428 asthma cases: 59 receiving and 369 not receiving biologic therapy; 271 controls) and blood DNAm data for 668 (408 asthma cases: 48 receiving and 360 not receiving biologic therapy; 260 controls) for cross-sectional analyses (**Table 1**). We evaluated 35 asthma PRS available from the Polygenic Score Catalog^2^, and developed six weighted MRS derived from EWAS of asthma in blood. An MRS derived from CpGs previously associated with atopic asthma in adults by Hoang et al.^3^ (MRS_adulthood-atopic-asthma_) was considered the primary MRS, as it aligns with GEMAS’s adult population and demonstrated high discriminatory performance. The remaining MRS and eosinophil-validated restricted versions of the Hoang^3^ and Reese^4^ scores were used in sensitivity analyses. We adjusted multiple comparisons using a false discovery rate (FDR) <0.05. Detailed methods are provided in the Supporting Information.

**Table 1.** Clinical and demographic characteristics of the study population (GEMAS).

| Characteristics | n | Healthy controls | n | Asthma without biologic | *p*-value vs control | n | Asthma with biologic | *p*-value vs control | *p*-value biologic vs non-biologic asthma |
| --- | --- | --- | --- | --- | --- | --- | --- | --- | --- |
| Sex (male) | 271 | 123 (45.4) | 369 | 134 (36.3) | **0.02** | 59 | 18 (30.5) | **0.04** | 0.39 |
| Age (years) | 271 | 44.0 (28.0-54.0) | 369 | 41.0 (28.0-54.0) | 0.44 | 59 | 51.0 (26.0-61.0) | 0.17 | 0.11 |
| BMI category, n (%) | 267 |  | 353 |  | **1.47x10<sup>-5</sup>** | 55 |  | **1.52x10<sup>-3</sup>** | 0.40 |
| Underweight |  | 5 (1.9) |  | 2 (0.6) |  |  | 0 (0.0) |  |  |
| Normal weight |  | 114 (42.7) |  | 107 (30.3) |  |  | 16 (29.1) |  |  |
| Overweight |  | 92 (34.5) |  | 126 (35.7) |  |  | 16 (29.1) |  |  |
| Obesity |  | 56 (21.0) |  | 118 (33.4) |  |  | 23 (41.8) |  |  |
| Asthma control, n (%) | - | - | 362 |  |  | 56 |  |  | 0.10 |
| Controlled |  | - |  | 108 (29.8) |  |  | 21 (35.6) |  |  |
| Partially controlled |  | - |  | 161 (44.5) |  |  | 18 (30.5) |  |  |
| Uncontrolled |  | - |  | 93 (25.7) |  |  | 17 (28.8) |  |  |
| Asthma severity, n (%) | - | - | 346 |  |  | 59 |  |  | **4.22x10<sup>-12</sup>** |
| Mild asthma |  | - |  | 52 (15.0) |  |  | 0 (0.0) |  |  |
| Moderate asthma |  | - |  | 21 (6.1) |  |  | 0 (0.0) |  |  |
| Severe asthma |  | - |  | 273 (78.9) |  |  | 59 (100) |  |  |
| Asthma exacerbations | - | - | 367 | 170 (46.3) |  | 59 | 32 (54.2) |  | 0.26 |
| Biologic therapy, n (%) | - | - | 369 | 0 (0.0) |  | 59 |  |  |  |
| Mepolizumab |  | - |  | - |  |  | 29 (49.2) |  |  |
| Omalizumab |  | - |  | - |  |  | 22 (37.3) |  |  |
| Dupilumab |  | - |  | - |  |  | 6 (10.2) |  |  |
| Benralizumab |  | - |  | - |  |  | 1 (1.7) |  |  |
| Reslizumab |  | - |  | - |  |  | 1 (1.7) |  |  |
| pre-FEV1, % predicted | - | - | 225 | 92.2 (77.3-103.0) |  | 46 | 84.7 (72.1-96.4) |  | 5.42x10<sup>-2</sup> |
| pre-FVC, % predicted | - | - | 225 | 97.0 (85.6-108.4) |  | 46 | 93.7 (80.7-107.9) |  | 0.52 |
| pre-FEV1/FVC, % | - | - | 225 | 95.2 (87.5-101.5) |  | 46 | 89.5 (83.9-99.6) |  | **4.90x10<sup>-2</sup>** |
| post-FEV1, % predicted | - | - | 77 | 95.5 (83.7-102.2) |  | 13 | 79.3 (75.8-87.9) |  | **5.67x10<sup>-3</sup>** |
| post-FVC, % predicted | - | - | 77 | 97.4 (90.2-107.9) |  | 13 | 91.1 (83.5-98.6) |  | 0.20 |
| post-FEV1/FVC, % | - | - | 77 | 95.0 (89.5-100.3) |  | 13 | 89.5 (83.0-92.1) |  | 1.70x10<sup>-2</sup> |
| tIgE | 136 | 116.0 (47.7-274.8) | 317 | 157.0 (55.1-523.0) | **4.64x10<sup>-2</sup>** | 56 | 262.5 (113.3-1327.0) | **1.10x10<sup>-4</sup>** | **5.85x10<sup>-3</sup>** |
| Blood eosinophils, cells/μl | 237 | 180.0 (100.0-310.0) | 341 | 300.0 (170.0-510.0) | **5.65x10<sup>-10</sup>** | 56 | 110.0 (42.5-337.5) | 0.07 | **3.15x10<sup>-5</sup>** |
| Blood eosinophils, % | 236 | 2.9 (1.8-4.6) | 338 | 4.4 (2.6-7.4) | **6.97x10<sup>-9</sup>** | 55 | 3.1 (1.0-7.4) | 0.99 | **3.89x10<sup>-2</sup>** |

Categorical variables are described as counts (percentage) and continuous variables as median (interquartile range). Exacerbations were defined by the occurrence of hospitalizations, emergency room visits, and/or oral corticosteroid use during the previous 12 months. Group comparisons were performed using Wilcoxon rank-sum tests for continuous variables and chi-square or Fisher's exact tests for categorical variables, as appropriate. Statistically significant differences are shown in bold (*p*-value<0.05). Body mass index categories were defined using age- and sex-specific BMI percentiles for participants <20 years and standard adult BMI cut-offs for participants ≥20 years: underweight, <18.5 kg/m<sup>2</sup>; normal weight, 18.5-24.9 kg/m<sup>2</sup>; overweight, 25.0-29.9 kg/m<sup>2</sup>; and obesity, ≥30.0 kg/m<sup>2</sup>. Lung function predicted values were calculated using the GLI 2022 reference equations. MRS analyses involving biologic-treated asthma were restricted to 48 patients receiving biologic therapy for ≥6 months. **Abbreviations:** BMI, body mass index; FEV<sub>1</sub>, forced expiratory volume in 1 second; FVC, forced vital capacity; tIgE, total immunoglobulin E.

Twenty-eight of the 35 asthma-annotated PRS differentiated biologic-treated asthma from healthy controls (**Figure S1**), AUCs=0.68-0.75. Although nine PRS nominally identified biologic-treated asthma as having higher genetic risk than non-biologic-treated asthma, none were significant after multiple-testing correction.

Regarding MRS, higher MRS_adulthood-atopic-asthma_ values were found in asthma without biologic versus controls (OR=1.64, 95% CI 1.34-1.99; *p*=8.68×10^−7^; **Figure 1A; Table S3**). Notably, patients receiving biologic therapy did not differ from controls (*p*=0.58) and had lower MRS_adulthood-atopic-asthma_ than individuals with asthma without biologic (OR=0.54, 95% CI 0.35-0.81; *p*=3.15×10^−3^; **Figure 1A; Table S3**). Among biologic-treated patients, anti-IL-5/IL-5R therapy was associated with lower MRS_adulthood-atopic-asthma_ than anti-IgE (*p*=0.013; **Figure 1B**).

**Figure 1.**
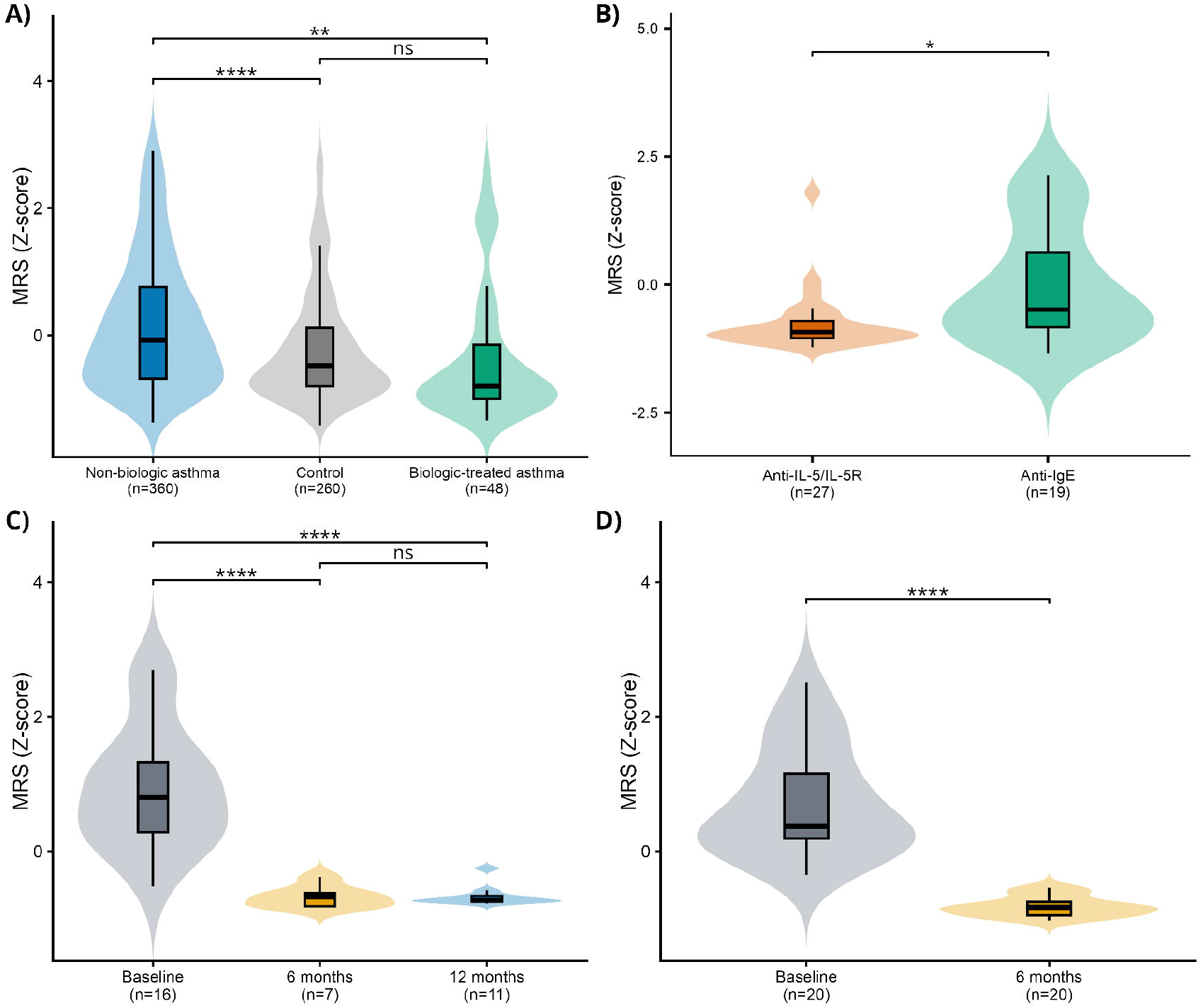
Cross-sectional and longitudinal variation in the Hoang et al. atopic asthma MRS. **(A)** Standardized MRS values in non-biologic asthma, controls, and biologic-treated asthma in GEMAS. **(B)** MRS values in biologic-treated patients according to biologic mechanism. **(C)** Longitudinal MRS values in GEMAS at baseline, 6 months, and 12 months after mepolizumab initiation. **(D)** Longitudinal MRS values in MEGA at baseline and 6 months. Violin plots show distributions, and boxplots show medians and interquartile ranges. Asterisks denote covariate-adjusted logistic regression *p*-values in A and B, and Tukey-adjusted linear mixed-model comparisons in C-D: \**p*<0.05, \*\**p*<0.01, \*\*\**p*<0.001, \*\*\*\**p*<0.0001; ns, not significant.

We next assessed longitudinal changes in MRS in 16 GEMAS patients with baseline and 6- and/or 12-month DNAm measurements after mepolizumab treatment (**Table S4**). The MRS_adulthood-atopic-asthma_ decreased at 6 months (adjusted mean difference=1.74; *p*=4.81×10^−5^) and 12 months (adjusted mean difference=1.51; *p*=2.40×10^−5^), with no evidence of further change between 6 and 12 months (*p*=0.80) (**Figure 1C; Table S5**). Most CpGs were significantly hypermethylated after treatment (92.1% at 6 months and 96.1% at 12 months) (https://doi.org/10.5281/zenodo.22870613), opposite to the predominant hypomethylation previously associated with asthma^3–5^. These results were replicated in 20 patients from the MEGA study (**Table S4**) with paired baseline and 6-month mepolizumab follow-up data (mean difference=1.56, SE=0.15; *p*=3.25×10^−9^; **Figure 1D**). Similar cross-sectional and longitudinal patterns were observed for the other evaluated MRSs, except for the score derived in African American and Hispanic/Latino populations^6^ (**Table S3; Table S5**), suggesting a low cross-ancestry portability of MRS. The longitudinal pattern also persisted in MRS eosinophil-restricted sensitivity analyses, based on CpGs previously associated with asthma in purified eosinophils (**Table S5**)^3,4^. This epigenetic reversal of asthma risk adds to reported molecular effects of mepolizumab^7,8^.

Strengths of this study include the evaluation of several asthma-related PRS, MRS derived from independent EWAS, longitudinal assessment, and replication. We acknowledge as limitations the predominant representation of European ancestry, modest longitudinal sample size, and the potential residual confounding from whole-blood cell composition in MRS results, despite including cell components as covariates in all models and performing eosinophil sensitivity analyses.

In conclusion, asthma patients treated with biologic therapy do not show large genetic differences compared with patients without biologic treatment, but they exhibit reduced epigenetic risk and a potential reversal of asthma-related DNAm patterns. PRS and MRS capture complementary aspects of asthma heterogeneity, supporting biologic treatment-related epigenetic remodeling.

## Data Availability

All data produced in the present work are contained in the manuscript and available online at https://doi.org/10.5281/zenodo.22870613

## Author contributions

MM-A, JP-G, RG-P, MP-Y were involved in the conceptualization of the study; MM-A, JP-G, and MP-Y in methodology; PP-G, JA-C, IS-M, JM-H, EM-L, BR-B, JV, MV, DB, MR-N, FL-D, VP, RG-P, and MP-Y in data acquisition and resources; MM-A, JP-G, EM-G, DG-G, and RG-P in data curation; MM-A in formal analysis and investigation; MM-A in visualization; MM-A, JP-G, and MP-Y in data interpretation and writing the original draft; RG-P and MP-Y in funding acquisition; and JP-G and MP-Y in project supervision. All authors were involved in critical revision and editing of the manuscript and agreed to the published version. The authors agree to be accountable for all aspects of the work, ensuring scientific accuracy and integrity.

## Acknowledgments

We would like to express our gratitude to all patients, families, recruiters, and participating clinical centers for their valuable contribution to the GEMAS and MEGA studies. The authors thank Centro Nacional de Genotipado□Plataforma de Recursos Biomoleculares□Instituto de Salud Carlos III (CeGen□PRB3□ISCIII; www.cegen.org) for providing genotyping services for the GEMAS study.

## Data Availability Statement

The data that support the findings of this study are openly available in Zenodo Repository at https://zenodo.org, reference number https://doi.org/10.5281/zenodo.22870613.

## Supporting information

Additional supporting information can be found online in the Supporting Information section. **Table S1**. Summary of the GEMAS genotype quality control results. **Table S2**. Summary of the GEMAS methylation quality control results. **Table S3**. Association and discrimination performance of asthma methylation risk scores across asthma-related comparisons. **Table S4**. Clinical characteristics of biologic-treated asthma patients at baseline and follow-up. **Table S5**. Longitudinal changes in all evaluated asthma methylation risk scores after mepolizumab therapy initiation. **Figure S1**. Forest plot of asthma-related PRS significantly associated with biologic-treated asthma versus controls at FDR<0.05 among the 35 asthma-annotated PRS evaluated.

