## Supporting Information for "Blood epigenetic remodeling in asthma associated with biologic therapy"

### Supplementary Methods

#### Study populations and study design

The main analyses included participants from the Genomics and Metagenomics of Asthma Severity (GEMAS) study^1^ recruited at Allergy and Respiratory Medicine units in hospitals across the Canary Islands. Eligible patients were aged 8-85 years and had physician-confirmed asthma. Individuals reporting non-European ancestry in at least one grandparent, along with first- or second-degree relatives, were not elegible. Controls had no asthma or other respiratory disease. To minimize potential phenotype misclassification, controls with records of biologic therapy, multiple allergic comorbidities, including atopy, rhinitis, or dermatitis, or a family history of asthma were excluded.

For PRS analyses, we analyzed 699 participants, including 271 controls and 428 patients with asthma, of whom 59 were receiving biologic therapy. Patients classified as asthma without biologic therapy had never received biologic treatment. Biologic-treated patients were receiving maintenance monoclonal antibody therapy for severe asthma at the time of blood sampling, including omalizumab, mepolizumab, benralizumab, reslizumab, or dupilumab. For MRS analyses, 668 participants were available after DNAm quality control, including 260 controls and 408 patients with asthma. The biologic-treated MRS group was restricted to 48 patients who had received biologic therapy for ≥6 months before blood sampling.

Longitudinal analyses included 16 GEMAS patients sampled before mepolizumab initiation and after 4-7 months (6-month follow-up, n=7) and/or 11-13 months (12-month follow-up, n=11). Two patients contributed to both follow-up windows. Replication was performed in 20 patients from the independent MEGA study^2^ with paired baseline and 6-month DNAm measurements. MEGA participants were adults with asthma recruited at three Spanish university hospitals and treated with mepolizumab.

All participants or their legal guardians provided written informed consent. The study was conducted in accordance with the Declaration of Helsinki and approved by the institutional review boards of all participating centers (Hospital Universitario de Canarias [29/17 and CHUC_2019_112], Hospital Universitario de Gran Canaria Doctor Negrín [2020-328-1], CHUC_BICMA [30-10-2025], and Fundación Jiménez Díaz [PIC002-19_FJD]).

#### Genotype data and polygenic risk scores

Participants were genotyped using the Illumina Global Screening Array v3.0. Sample and variant-level quality control included filtering based on genotype call rate, deviation from Hardy–Weinberg equilibrium, sex discordance, heterozygosity, and relatedness. Principal component analysis was performed to identify ancestry outliers. After quality control, 699 participants and 545,765 variants were retained. Genotypes were imputed, and PRS were calculated through the Michigan Imputation Server^3^ using published PRS from the Polygenic Score Catalog^4^. Among 4,489 available scores, ten were excluded because of unavailable SNPs or unspecified target ancestry, leaving 4,479 successfully calculated PRS representing >580 traits. Of these, 35 PRS with a target trait annotated as asthma in the PGS Catalog were selected for the analyses. PRS values were standardized to Z-scores before analysis.

#### DNA methylation and methylation risk scores

Whole-blood DNAm was profiled using the Illumina Infinium MethylationEPICv1 and v2 arrays in GEMAS and EPICv2 in MEGA. Quality control and normalization were performed separately by array version using the *ENmix*^5^ and *ewastools*^6^ R packages, with similar procedures as described elsewhere^7^. Samples with poor overall methylation quality, sex discordance, or evidence of cross-sample contamination were excluded. Probe-level filtering removed low-quality, cross-reactive, polymorphic, sex-chromosome, and array-specific problematic probes. Estimated blood cell proportions were obtained using the Houseman reference-based approach^8^.

After quality control, GEMAS EPIC v1 and EPIC v2 datasets were harmonized by retaining CpGs common to both platforms. Technical variation related to array version and experimental batch was corrected on beta values using ComBat-met^9^. A total of 633,569 CpGs from 668 GEMAS participants were retained. Longitudinal GEMAS analyses were restricted to EPIC v2 data and therefore did not require cross-platform harmonization. MEGA was also profiled using EPIC v2, retaining 787,367 CpGs and 40 samples from 20 participants with paired baseline and 6-month measurements.

We constructed six *de novo* asthma MRS based on published blood EWAS results, and evaluated them for asthma case-control associations. Hoang et al.^10^ identified 181 (145 available in GEMAS) and 8 (6 available) epigenome-wide significant CpGs for atopic and non-atopic asthma, respectively, in European adults. Pediatric MRS included the 179 asthma-associated CpGs identified in the childhood meta-EWAS by Reese et al.^11^ (140 available), the 26 CpGs identified in the discovery analysis by Xu et al.^12^ (20 available), and 7 CpGs from an independent replication meta-analysis reported by Xu et al. (4 available). An additional MRS was derived from the multi-ancestry childhood EWAS by Herrera-Luis et al.^13^ in African American and Hispanic/Latino participants, with 104/114 CpGs available. In the longitudinal GEMAS dataset, 152 CpGs were available for the Hoang et al. atopic asthma MRS, 6 for the Hoang et al. non-atopic asthma MRS, 148 for Reese et al., 21 for Xu et al., and 4 for the Xu et al. replication meta-analysis. The Herrera-Luis et al. MRS was evaluated cross-sectionally only. Each MRS was calculated as the weighted sum of CpG beta values using the published effect estimates and standardized to a Z-score.

#### Statistical analyses

PRS and MRS associations were evaluated using logistic regressions. PRS models were adjusted for age, sex, and the first two genetic principal components (PCs). MRS models were additionally adjusted for estimated blood cell proportions. Odds ratios (ORs) were reported per 1-SD increase in each score. Model discrimination was assessed using the area under the receiver operating characteristic curve (AUC).

For PRS, two cross-sectional comparisons were evaluated: 1) asthma with biologic therapy versus controls and 2) asthma with biologic therapy versus without biologic therapy. PRS analyses were restricted to the 35 PRS annotated for asthma in the PGS Catalog, with FDR correction performed across these scores within each comparison. For MRS, four cross-sectional comparisons were evaluated: 1) asthma versus controls, 2) asthma without biologic therapy versus controls, 3) asthma with biologic therapy versus controls, and 4) asthma with biologic therapy versus without biologic therapy. FDR correction was performed across the six MRS within each comparison. Statistical significance was defined as FDR<0.05. Among biologic-treated patients, the association between MRS and biologic mechanism (anti-IgE versus anti-IL-5/IL-5R) was evaluated using logistic regression with the same covariate adjustments. Patients receiving anti-IL-4/IL-13 therapy were excluded from this comparison because only two had DNA methylation data.

Longitudinal MRS changes were assessed using linear mixed-effects models with a participant-specific random intercept to account for repeated measurements. Models were adjusted for age, sex, genetic PCs, and cell proportions in GEMAS. The same approach was applied in MEGA, except that genetic PCs were not included because genotype data were unavailable. Pairwise contrasts were evaluated between baseline and 6 months, baseline and 12 months, and 6 and 12 months in GEMAS, and between baseline and 6 months in MEGA. FDR correction was performed across the five primary longitudinal MRS within each timepoint comparison.

Longitudinal CpG-level changes were evaluated separately within each MRS CpG set using linear models implemented in *limma*^14^, with within-subject correlation estimated using duplicateCorrelation and participant ID included as a blocking factor. Models were adjusted for the same covariates as longitudinal MRS analyses. FDR correction was recalculated within each CpG set and follow-up window. The direction of post-treatment DNAm changes was compared with the asthma-associated effect reported in the original EWAS to determine whether methylation changes after mepolizumab occurred or not in the opposite direction to the asthma-associated pattern.

#### Eosinophil-related sensitivity analysis

Given the marked eosinophil depletion induced by mepolizumab, we performed sensitivity analyses using eosinophil-restricted versions of the Reese et al. and Hoang et al. MRS^10,11^. In Reese et al., 177 of 179 asthma-associated CpGs were available in purified eosinophils, all with concordant directions, and 148 were associated with asthma at *p*<0.05. In Hoang et al., among 178 genome-wide significant atopic asthma CpGs, 49 replicated in purified eosinophils at *p*<0.05. We reconstructed both MRS using the eosinophil-replicated CpGs available after DNAm quality control in the longitudinal GEMAS dataset (Reese: 123 CpGs; Hoang: 42 CpGs), weighted by the original whole-blood EWAS effect estimates. Longitudinal models were then repeated using these scores.

### Supplementary Figures and Tables

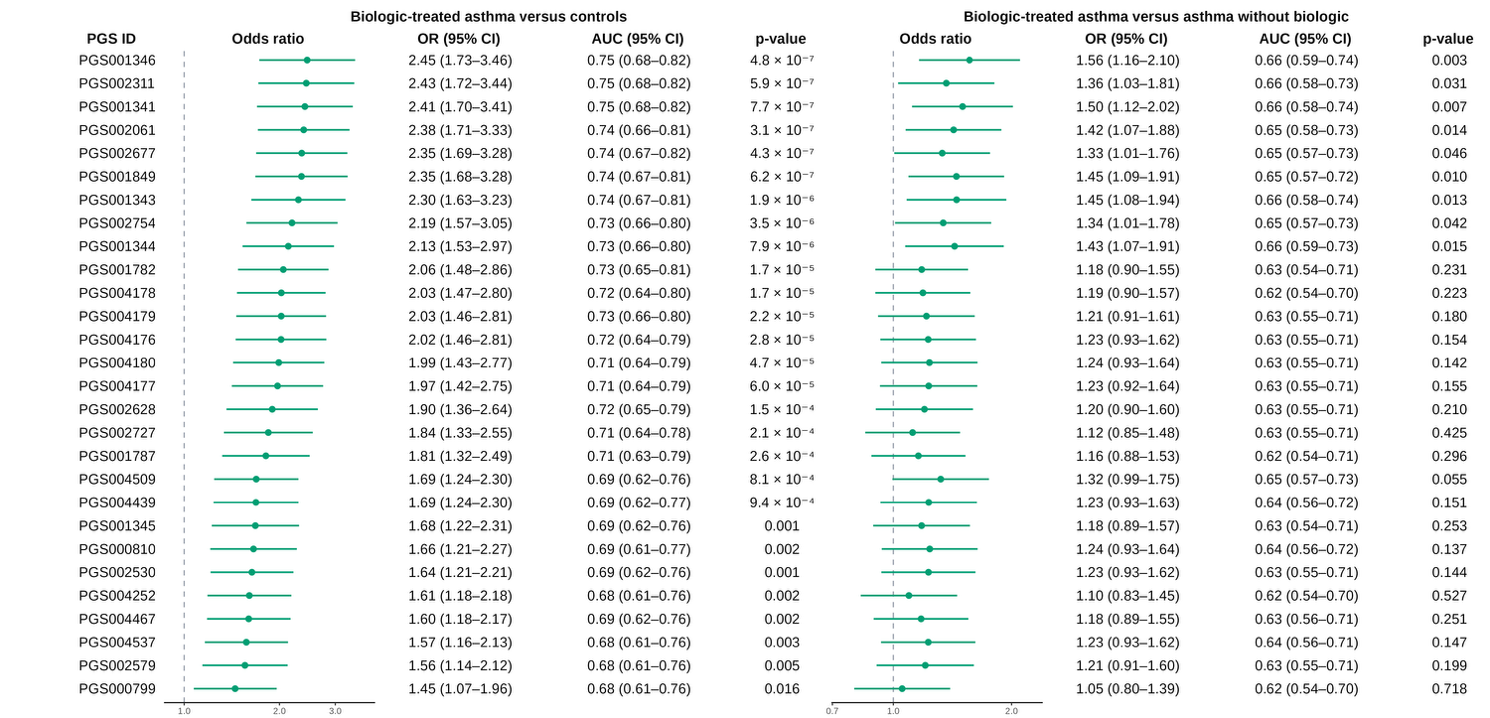

**Supplementary Figure 1.** Forest plot of asthma-related polygenic risk scores (PRS) significantly associated with biologic-treated asthma versus controls at FDR<0.05 among the 35 asthma-annotated PRS evaluated. For these PRS, associations are shown for biologic-treated asthma versus controls and biologic-treated versus non-biologic asthma. Points represent adjusted odds ratios (OR) per one-standard-deviation increase in PRS, and horizontal bars indicate 95% confidence intervals (CI). Models were adjusted for age, sex, and the first two genetic principal components. Vertical dashed lines denote the null value (OR=1). Corresponding OR estimates, 95% CI, and *p*-values are displayed. PGS IDs correspond to identifiers in the Polygenic Score Catalog.

| **Supplementary Table 1**. Summary of the GEMAS genotype quality control results. | |
| --- | --- |
| **Quality control of variants** | **n** |
| Initial number of variants | 650,181 |
| Variants not assigned to chromosomes | 1,211 |
| Monomorphic variants | 98,879 |
| Duplicated variants | 2,167 |
| Variants without rsID | 107 |
| Variants with deviations from HWE in controls | 108 |
| Variants with call rate <95% | 1,944 |
| Total remaining genetic variants | 545,765 |
| **Quality control and exclusion criteria for individuals** | |
| Initial number of individuals | 892 |
| Individuals with sex discordance | 4 |
| Individuals with missing genotype (call rate < 95%) | 12 |
| Related individuals (PIHAT≥0.25) | 17 |
| Duplicated individuals | 23 |
| Individuals recruited in the Basque country* | 109 |
| Ancestry outliers identified by PCA | 17 |
| Controls excluded based on clinical criteria | 11 |
| Total remaining samples | 699 |
| *Individuals recruited in the Basque Country were excluded because this recruitment site included only asthma cases. **Abbreviations:** rsID, reference SNP identifier; HWE, Hardy-Weinberg equilibrium; PCA, principal component analysis; PIHAT, proportion of identity by descent. Controls excluded based on clinical criteria included individuals with records of biologic therapy, multiple allergic comorbidities, or a family history of asthma. | |

| **Supplementary Table 2.** Summary of the GEMAS methylation quality control results. | | |
| --- | --- | --- |
| Array | **EPIC v1** | **EPIC v2** |
| **Quality control of samples** |  |  |
| Initial number of individuals | 476 | 267 |
| Did not pass genotype quality control | 9 | 0 |
| Outlier or bad quality samples | 4 | 0 |
| Individuals with missingness >10% | 4 | 1 |
| Individuals with mixed genotype distributions on the control SNP probes | 4 | 0 |
| PCA outliers | 6 | 0 |
| Individuals with sex discordance | 1 | 0 |
| Total remaining samples | 448 | 266 |
| **Quality control of CpG sites** |  |  |
| Initial number of CpGs | 865,918 | 937,055 |
| **Removed probes** |  |  |
| Bad quality probes (detection *p*-value) | 25,666 | 5,993 |
| Bad quality probes (missing data across samples) | 15,182 | 5,487 |
| Flagged Illumina probes | 826 | 57,406 |
| Cancer-associated variant probes (nv) | - | 824 |
| Not annotated probes | - | 7,551 |
| Replicate probes | - | 4,993 |
| Multimodal probes | 7,238 | 5,481 |
| Cross-reactive probes^15–19^ | 83,684 | 40,303 |
| Control SNP genotyping probes | 57 | 54 |
| SNP at a CpG site | 7,186 | 8,886 |
| SNP at SBE from a CpG site | 3,570 | 3,692 |
| Probes in sex chromosomes | 18,627 | 24,509 |
| Bad probes in positive control (β<0.8) | 5,405 | - |
| Bad probes in an EPICv1 technical duplicate (Δβ≥0.2)* | 2,368 | - |
| Total remaining CpGs | 703,505 | 806,132 |
|  | **EPICv1 + EPICv2** | |
| Initial number of individuals | 714 | |
| Follow-up samples retained for longitudinal analyses | 22 | |
| Duplicated or related individuals | 14 | |
| Controls excluded based on clinical criteria | 10 | |
| Samples for cross-sectional analyses | 668 (260 controls and 408 cases) | |
| Final number of CpG sites analyzed (overlapping between EPICv1 + EPICv2) | 633,569 | |
| **Abbreviations.** CpG, cytosine-phosphate-guanine site; PCA, principal component analysis; SNP, single nucleotide polymorphism. Controls excluded based on clinical criteria included individuals with records of biologic therapy, multiple allergic comorbidities, or a family history of asthma. Follow-up samples were retained for longitudinal analyses but were not included in the primary cross-sectional analyses. The final cross-sectional sample size refers to unrelated baseline samples. *One sample was assayed in duplicate on EPICv1 to assess technical concordance. Probes showing Δβ≥0.2 between duplicate measurements were excluded. | | |

| **Supplementary Table 3.** Association and discrimination performance of asthma methylation risk scores across asthma-related comparisons. | | | | | |
| --- | --- | --- | --- | --- | --- |
| Reference study for MRS estimation | **Comparison** | **OR (95% CI)** | **AUC (95% CI)** | ***p*-value** | **FDR** |
| Hoang et al.^10^ atopic asthma  (MRS_adulthood-atopic-asthma_) | Asthma vs control | 1.53 (1.27-1.84) | 0.70 (0.66-0.74) | 8.40x10^-6^ | 1.26x10^-5^ |
|  | Asthma without biologic vs control | 1.64 (1.34-1.99) | 0.72 (0.68-0.76) | 8.68x10^-7^ | 1.30x10^-6^ |
|  | Asthma with biologic vs control | 0.89 (0.60-1.33) | - | 0.58 | 0.83 |
|  | Asthma with biologic vs asthma without biologic | 0.54 (0.35-0.81) | 0.74 (0.66-0.82) | 3.15x10^-3^ | 5.18x10^-3^ |
| Hoang et al.^10^ non atopic asthma  (MRS_adulthood-non-atopic-asthma_) | Asthma vs control | 1.62 (1.35-1.96) | 0.70 (0.66-0.74) | 4.12x10^-7^ | 2.47x10^-6^ |
|  | Asthma without biologic vs control | 1.75 (1.43-2.13) | 0.72 (0.68-0.76) | 3.98x10^-8^ | 2.39x10^-7^ |
|  | Asthma with biologic vs control | 0.96 (0.66-1.40) | - | 0.85 | 0.85 |
|  | Asthma with biologic vs asthma without biologic | 0.57 (0.39-0.84) | 0.73 (0.65-0.81) | 4.32x10^-3^ | 5.18x10^-3^ |
| Reese et al.^11^  (MRS_childhood-asthma-Reese_) | Asthma vs control | 1.55 (1.29-1.87) | 0.70 (0.66-0.74) | 3.74x10^-6^ | 7.48x10^-6^ |
|  | Asthma without biologic vs control | 1.66 (1.37-2.02) | 0.72 (0.68-0.76) | 3.33x10^-7^ | 6.66x10^-7^ |
|  | Asthma with biologic vs control | 0.92 (0.63-1.36) | - | 0.69 | 0.83 |
|  | Asthma with biologic vs asthma without biologic | 0.55 (0.37-0.82) | 0.74 (0.66-0.82) | 3.73x10^-3^ | 5.18x10^-3^ |
| Xu et al.^12^  (MRS_childhood-asthma-Xu_) | Asthma vs control | 1.48 (1.23-1.78) | 0.69 (0.65-0.73) | 3.81x10^-5^ | 4.57x10^-5^ |
|  | Asthma without biologic vs control | 1.59 (1.31-1.93) | 0.71 (0.67-0.75) | 3.78x10^-6^ | 4.54x10^-6^ |
|  | Asthma with biologic vs control | 0.85 (0.57-1.26) | - | 0.41 | 0.83 |
|  | Asthma with biologic vs asthma without biologic | 0.53 (0.36-0.80) | 0.74 (0.66-0.82) | 3.73x10^-3^ | 5.18x10^-3^ |
| Xu et al.^12^ meta-analysis  (MRS_childhood-asthma-Xu-replication_) | Asthma vs control | 1.57 (1.30-1.90) | 0.70 (0.66-0.74) | 2.39x10^-6^ | 7.17x10^-6^ |
|  | Asthma without biologic vs control | 1.70 (1.39-2.07) | 0.72 (0.68-0.76) | 1.72x10^-7^ | 5.16x10^-7^ |
|  | Asthma with biologic vs control | 0.91 (0.61-1.34) | - | 0.61 | 0.83 |
|  | Asthma with biologic vs asthma without biologic | 0.50 (0.33-0.77) | 0.74 (0.66-0.82) | 1.37x10^-3^ | 5.18x10^-3^ |
| Herrera-Luis et al.^13^  (MRS_childhood-asthma-Herrera_) | Asthma vs control | - | - | 0.34 | 0.34 |
|  | Asthma without biologic vs control | - | - | 0.33 | 0.33 |
|  | Asthma with biologic vs control | - | - | 0.60 | 0.83 |
|  | Asthma with biologic vs asthma without biologic | - | - | 0.49 | 0.49 |
| Reported are ORs per 1 SD increase in MRS, AUCs, 95% CIs, and *p*-values from adjusted logistic regression models including age, sex, PC1-PC2, and Houseman cell-type proportions. FDR correction was applied across the six MRSs evaluated within each comparison. ORs and full-model AUCs are shown only for MRS associations passing FDR<0.05; non-significant associations are indicated by “-”. | | | | | |

| **Supplementary Table 4.** Clinical characteristics of biologic-treated asthma patients at baseline and follow-up. | | | | | | | | | |
| --- | --- | --- | --- | --- | --- | --- | --- | --- | --- |
|  | **GEMAS, 6 months (n=7)** | | | **GEMAS, 12 months (n=11)** | | | **MEGA, 6 months (n=20)** | | |
|  | **Baseline** | **Follow-up** | ***p*-value** | **Baseline** | **Follow-up** | ***p*-value** | **Baseline** | **Follow-up** | ***p*-value** |
| Sex (female) | 5 (71.4) | | - | 7 (63.6) | | - | 14 (70.0) | | - |
| Age (years) | 49.0  (37.5 - 51.5) | 50.0  (38.0 - 52.0) | - | 50.0  (48.0 - 58.0) | 51.0  (49.0 - 58.5) | - | 62.5  (54.3 - 69.0) | 63.0  (54.9 - 69.6) | - |
| BMI (kg/m^2^) | 33 (26.1 - 34.9) | | - | 30.1 (26.9 - 35.9) | | - | 24.3 (20.6 - 28.0) | | - |
| Eosinophils (cells/$\mu l)$ | 700.0  (470.0 - 725.0) | 50.0  (47.5 - 70.0) | **0.02** | 590  (415.0 - 665.0) | 70  (40.0 - 90.0) | **4.0x10^-3^** | 550.0  (425.0 - 650.0) | 50.0  (0.0 - 100.0) | **0.01** |
| IgE (IU/mL) | 163.0  (148.5 - 306.9) | 136  (118.7 - 351.0) | 0.8 | 137  (60.4 - 271.0) | 79.8  (54.9 - 144.6) | 0.08 | 736.0  (233.0 - 1460.0) | 683.0  (191.0 - 1187.0) | 0.53 |
| Exacerbations | 3  (2.0 - 4.5) | 0  (0.0 - 1.5) | **0.02** | 2  (1.5 - 2.5) | 0  (0.0 - 1.0) | **5.0x10^-3^** | 3.0  (3.0 - 4.75) | 0  (0.0 - 0.0) | **5.5x10^-3^** |
| ER | 3  (2.5 - 4.5) | 0  (0.0 - 1.0) | **0.02** | 2.0  (1.5 - 2.5) | 0  (0.0 - 1.0) | **8.0x10^-3^** | - | - | - |
| HOSP | 0  (0.0 - 2.0) | 0  (0.0 - 0.5) | 0.37 | 0.0  (0.0 - 0.0) | 0  (0.0 - 0.0) | 1 | - | - | - |
| OCS | 3  (2.5 - 4.5) | 0  (0.0 - 1.5) | **0.02** | 2.0  (1.5 - 3.0) | 0  (0.0 - 0.0) | **3.0x10^-3^** | - | - | - |
| FEV_1_ (%, predicted) | 85.3  (77.2 - 88.2) | 79.2  (64.7 - 84.7) | 1 | 85.9  (63.7 - 104.7) | 80.0  (62.7 - 92.1) | 1 | 95.7  (68.7 - 105.1) | 102.5  (97.3 - 105.8) | 0.10 |
| FeNO (ppb) | 65  (26.5 - 101.5) | 72  (23.5 - 117.0) | 0.7 | 39  (18.0 - 64.5) | 32  (26.0 - 59.0) | 0.48 | 22.7  (16.2 - 63.3) | 45.4  (26.4 - 67.7) | 1.00 |
| Categorical variables are described as counts (percentage) and continuous variables as median (interquartile range). Baseline and follow-up values correspond to measurements at treatment initiation and follow-up windows. Paired comparisons between baseline and recontact were performed using paired t-tests or Wilcoxon signed-rank tests, as appropriate. Statistically significant differences are shown in bold (*p*-value<0.05). Exacerbations, emergency room visits, hospitalizations, and oral corticosteroid courses refer to events occurring during the previous 12 months. **Abbreviations:** tIgE, total immunoglobulin E; ER, emergency room; HOSP, hospitalization; OCS, oral corticosteroids; BMI, body mass index; FeNO, fractional exhaled nitric oxide. | | | | | | | | | |

| **Supplementary Table 5**. Longitudinal changes in all evaluated asthma methylation risk scores after mepolizumab therapy initiation. | | | | | | |
| --- | --- | --- | --- | --- | --- | --- |
| **MRS** | **Comparison** | **Pairs** | **Adjusted mean difference** | **Standard Error** | ***p*-value** | **FDR *p*-value** |
| Hoang et al.^10^ atopic asthma  (MRS_adulthood-atopic-asthma_) | Baseline vs 6 months | 7 | 1.74 | 0.30 | 4.81x10^-5^ | 6.01x10^-5^ |
|  | Baseline vs 12 months | 11 | 1.51 | 0.24 | 2.40x10^-5^ | 3.00x10^-5^ |
|  | 6 months vs 12 months | 2 | -0.23 | 0.36 | 0.80 | 0.95 |
| Hoang et al.^10^ non atopic asthma  (MRS_adulthood-non-atopic-asthma_) | Baseline vs 6 months | 7 | 1.49 | 0.31 | 3.34x10^-4^ | 3.34x10^-4^ |
|  | Baseline vs 12 months | 11 | 1.38 | 0.24 | 7.54x10^-5^ | 7.54x10^-5^ |
|  | 6 months vs 12 months | 2 | -0.11 | 0.36 | 0.95 | 0.95 |
| Reese et al.^11^  (MRS_childhood-asthma-Reese_) | Baseline vs 6 months | 7 | 1.79 | 0.30 | 3.01x10^-5^ | 6.01x10^-5^ |
|  | Baseline vs 12 months | 11 | 1.51 | 0.24 | 2.16x10^-5^ | 3.00x10^-5^ |
|  | 6 months vs 12 months | 2 | -0.28 | 0.35 | 0.71 | 0.95 |
| Xu et al.^12^  (MRS_childhood-asthma-Xu_) | Baseline vs 6 months | 7 | 1.72 | 0.30 | 3.99x10^-5^ | 6.01x10^-5^ |
|  | Baseline vs 12 months | 11 | 1.52 | 0.24 | 1.69x10^-5^ | 3.00x10^-5^ |
|  | 6 months vs 12 months | 2 | -0.21 | 0.35 | 0.83 | 0.95 |
| Xu et al.^12^ meta-analysis  (MRS_childhood-asthma-Xu-replication_) | Baseline vs 6 months | 7 | 1.78 | 0.30 | 2.78x10^-5^ | 6.01x10^-5^ |
|  | Baseline vs 12 months | 11 | 1.51 | 0.24 | 1.85x10^-5^ | 3.00x10^-5^ |
|  | 6 months vs 12 months | 2 | -0.27 | 0.35 | 0.73 | 0.95 |
| Reese et al.^11^ eosinophils sensitivity analysis | Baseline vs 6 months | 7 | 1.75 | 0.29 | 2.75x10^-5^ | - |
|  | Baseline vs 12 months | 11 | 1.47 | 0.23 | 1.99x10^-5^ | - |
|  | 6 months vs 12 months | 2 | -0.28 | 0.34 | 0.70 | - |
| “Pairs” indicates the number of participants included in each paired comparison. Adjusted mean differences represent estimated differences in standardized MRS between time windows after accounting for age, sex, PC1-PC2, and estimated cell-type proportions. Subject was included as a random intercept to account for repeated measurements from the same participant. Positive adjusted mean differences indicate higher MRS values in the first-time window than in the second. Standard errors and *p*-values were obtained from the linear mixed model. FDR<0.05 correction was applied across the five methylation risk scores evaluated within each longitudinal comparison. | | | | | | |
